# Patterns of West Nile virus in the Northeastern United States using negative binomial and mechanistic trait-based models

**DOI:** 10.1101/2022.11.09.22282143

**Authors:** Alexander C. Keyel

## Abstract

West Nile virus primarily infects birds and mosquitoes but has also caused over 2000 human deaths, and >50,000 reported human cases. Present-day West Nile virus risk was described for the Northeastern United States, using a negative binomial model. Changes in risk due to climate change were examined for the next decade using a temperature-trait model. West Nile virus risk was generally expected to increase over the next decade due to changes in temperature, but the changes in risk were generally small. Many, but not all, populous counties in the northeast are already near peak risk. Several years in a row of low case numbers is consistent with a negative binomial, and should not be interpreted as a change in disease dynamics. Public health budgets need to be prepared for the expected infrequent years with higher-than-average cases. Absence of reported cases from low-population counties is consistent with similar risk as nearby counties with cases.

## Introduction

Climate change is predicted to adversely affect human health and economic productivity (USGCRP, 2018, p. 18). One way climate change is expected to affect human health is through changes to patterns of infectious disease (e.g., Ryan et al., 2015). West Nile virus (WNV) is a vector-borne disease of public health concern (Hayes et al., 2005; Keyel, Gorris, et al., 2021) and is expected to change its distribution due to climate change (Chen et al., 2013; Hoover & Barker, 2016; Keyel, Raghavendra, et al., 2021). Broadly, WNV is expected to shift northward, but regional temperature-based analyses show that changes may vary depending on regional differences in temperature (Keyel, Raghavendra, et al., 2021; Morin & Comrie, 2013). For most of the Northeast, temperatures are predicted to warm, especially minimum (night-time) temperatures (Liu et al., 2017). Precipitation is also predicted to increase, especially in winter, due in part to an increased number of storms (Lynch et al., 2016; Thibeault & Seth, 2014). Summers may see increased run-off and periods of dryness (Lynch et al., 2016). In the next ten years, the climate is expected to warm by 0.2 – 0.5 °C (Liu et al., 2017). These predicted changes are within the range of variation in temperature currently experienced (Fig. 1).

**Fig. 1.**
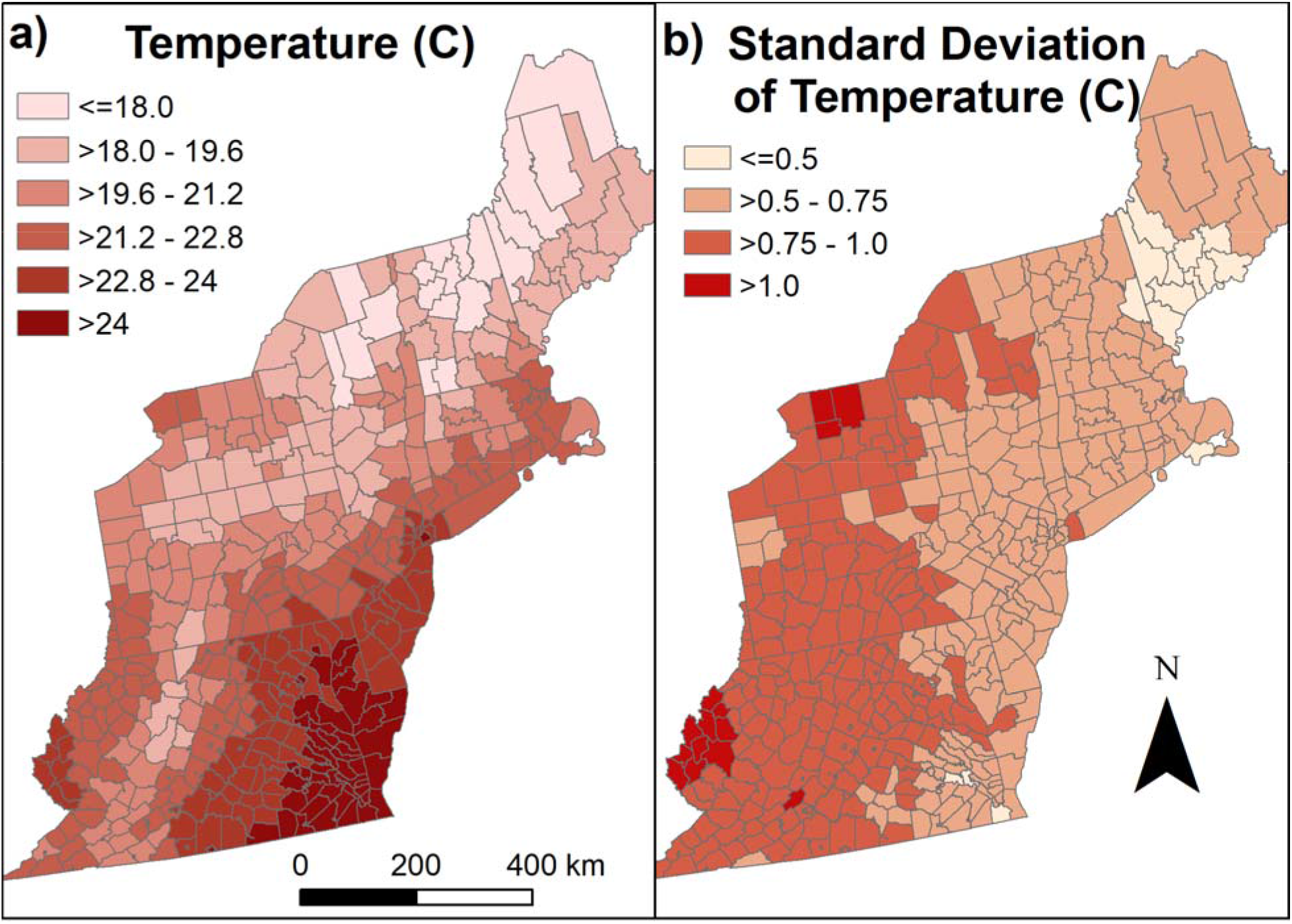
a) Present-day mean temperature for July - September, b) standard deviation of present-day temperature. Map base layer from 2017 TIGER/Shapefiles (US Census Bureau, 2021).

Probabilistic null models have been previously developed for the United States (Keyel et al., unpublished). These models consider a range of possible outcomes, rather than predicting one single number of cases for the future. In one instance, a model that worked well in a non-probabilistic context (e.g., Keyel et al., 2019) was not able to outperform a probabilistic negative binomial null model in a predictive context. As a consequence, current models for the Northeastern US are very good at describing the range of possible outcomes, but do not provide much information on where in the range of outcomes a particular year will fall.

The goal of this manuscript was to apply the probabilistic null models to gain useful insights into patterns of West Nile virus, and compare them to short-range climate change predictions with an aim of identifying short-term adaptation measures that can be taken.

## Methods

### Negative binomial model

Negative binomial model predictions were taken from Keyel et al. (Keyel et al., unpublished), as that was found to be among the strongest null models in the Northeast. The negative binomial was chosen instead of the historical null model due to the long time series, and the capacity to downscale model results based on population (Klenke, 2008). The model used here was implemented in R (R Core Team, 2017). This distribution allowed us to cleanly calculate probability of an arbitrary number of human neuroinvasive cases for an arbitrary number of years into the future. While this model does not identify a good year vs. a bad year, it can give insights into the overall probability of a bad year for WNV.

A negative binomial was fit to each county individually, and to groups of multiple counties (Figure S1). Group assignment was subjective and followed the following guidelines: Each group needed to be contiguous with surrounding counties, contain counties from only one state, and include a minimum of 600,000 people in each group. No upper group size was imposed, but groups with more than 1,200,000 were examined to see if they could be split into two or more groups. Secondarily, counties with similar population density were preferentially grouped together. The probability a county would have zero West Nile virus cases by chance was calculated, assuming the down-scaled group negative binomial distribution was true.

Neuroinvasive case data are available from the Centers for Disease Control ArboNET (CDC, 2019), and R code is available from a public repository (software citation pending). Maps were created using ArcGIS 10.6.1 (Redlands, CA) with 2017 US Census TIGER/Shapefiles as the county base layer (US Census Bureau, 2021). Population data were derived from US Census data (US Census Bureau, 2017, 2019).

### Climate change predictions

Mosquito-temperature-trait models have been productively used to understand vector-borne diseases (Mordecai et al., 2019). They were recently adapted to WNV (Shocket et al., 2020). It was previously found that statistical models trained on human cases that included an adequate temperature range largely supported the results of mosquito-trait-based models for West Nile virus in New York and Connecticut (Keyel, Raghavendra, et al., 2021). This manuscript builds upon this prior work by expanding the use of the mosquito temperature-trait models to the entire northeast. Briefly, these models use mosquito life history traits to estimate a relative R_0_ for mosquito risk based on temperature. Traits are combined in a multiplicative framework (see Eqn 1, modified from (Shocket et al., 2020), see (Keyel, Raghavendra, et al., 2021) for details), then scaled to give an index of risk between 0 and 1.

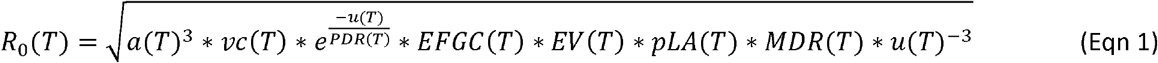

Host density and disease duration were assumed to be constant (but see Kilpatrick et al., 2006 for the potential for host heterogeneity to affect these results). Critically, these models look at the contribution of temperature only to risk, other important factors such as breeding habitat availability, land cover (Bradley et al., 2008), or precipitation are not considered (Shocket et al., 2020). Please note that these models were developed in the context of *Culex pipiens*, and therefore predicted risk will likely be different for different mosquito species (Shocket et al., 2020). However, *Cx. pipiens* is one of the most important mosquito vectors across the Northeastern US (Andreadis, 2012; Kilpatrick et al., 2005; Simpson et al., 2012; Turell et al., 2005). Present day conditions and risk expected for 0.5 °C warming were examined. Present-day July – September mean temperatures were derived from gridMET (Abatzoglou, 2013), using the GridMET downloader tool (Wimberly & Davis, 2019) and averaged over the entire period. Model results up to 4°C warming were generated and are available (data citation pending). Up to 4°C warming for the 5 most densely populated counties in the Northeast was also examined (Table 1). Warming of up to 4°C is within the realm of possible temperature changes for the Northeast by the end of the century (IPCC, 2014; Liu et al., 2017). This is also the rationale for 0.5°C for an upper-bound for the increase in temperature in the next decade (4°C / 8 decades = 0.5°C per decade). Note that the analyses here calculated trait-based risk on mean temperatures for the region. Due to the non-linear response curve, the quantitative results would have differed if relative risk were calculated first, and then averaged. A second source of error for this approach is microclimatic variability, which can affect disease risk (Haider et al., 2017). Future research could refine these results, but the broad patterns are expected to be qualitatively similar.

**Table 1.** Relative R_0_’s for 5 major northeastern US metropolitan counties based on different levels ofwarming (in C).

| County | +0.0 | +0.5 | +1.0 | +1.5 | +2.0 | +2.5 | +3.0 | +3.5 | +4.0 |
| --- | --- | --- | --- | --- | --- | --- | --- | --- | --- |
| Suffolk (Boston) | 0.88 | 0.92 | 0.96 | 0.99 | 1.00 | 1.00 | 0.98 | 0.96 | 0.92 |
| New York (Manhattan) | 1.00 | 1.00 | 0.98 | 0.95 | 0.91 | 0.86 | 0.80 | 0.73 | 0.66 |
| Philadelphia | 1.00 | 1.00 | 0.99 | 0.96 | 0.92 | 0.88 | 0.82 | 0.75 | 0.68 |
| Baltimore City | 0.98 | 0.95 | 0.91 | 0.86 | 0.80 | 0.74 | 0.66 | 0.58 | 0.50 |
| District of Columbia | 1.00 | 0.99 | 0.96 | 0.92 | 0.87 | 0.82 | 0.75 | 0.67 | 0.597 |

## Results

Most counties (374 of 433) in the study region had a low probability (<20% chance) of having a single WNV neuroinvasive case in the next year (Fig. 2a). More than three times more counties had a high risk of having at least one neuroinvasive case (>60% chance) in a five-year time frame (63, Fig. 2b) compared to a 1-year timeframe (17 of 433, Fig. 2a). Relatively few counties (10 of 433) had a high probability (>60% chance) of having a year with at least 5 neuroinvasive cases within a 5-year timeframe, and these were predominantly urban areas with high populations (Fig. 2c). Two groups, containing 40 counties, never had a single case of WNV. For the remaining groups with WNV cases, counties with no observed cases were predicted to have a low probability of cases based on the group model. Only one county (Washington, PA) had a <0.05 probability of having 20 years of no cases by chance assuming a similar risk to the rest of the counties in the group. When corrected for multiple comparisons, no county differed significantly from the negative binomial model (with 174 comparisons, ∼8-9 counties would be expected to have a p value <0.05 by chance).

**Fig. 2.**
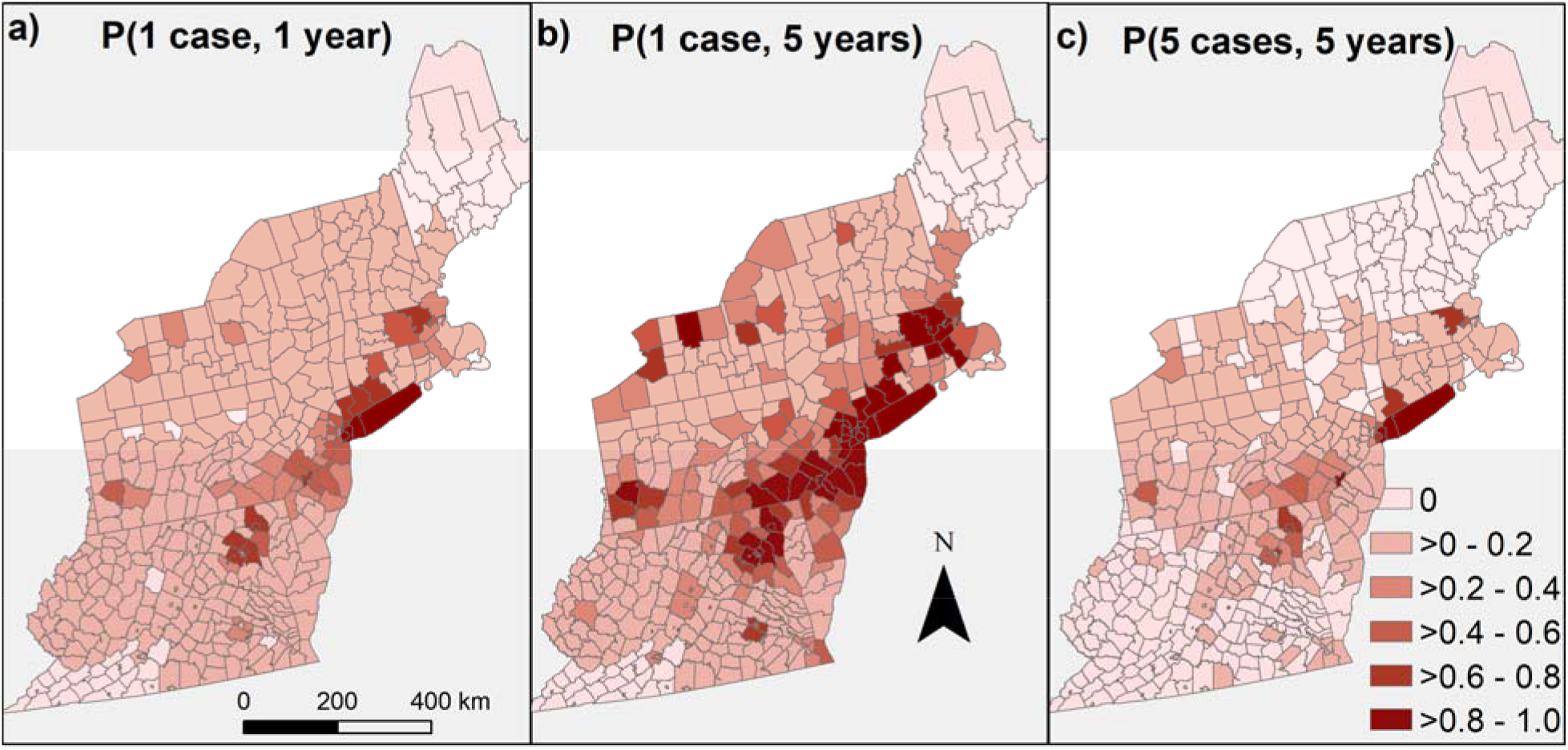
a) Probability of having one case of West Nile virus in the next year, b) probability of having at least one case of WNV in the next five years, and c) probability of having one year with at least 5 cases in the next five years. The negative binomial model was fit based on historical cases. Counties with less than 600,000 people were merged with other contiguous counties in the same state until at least a 600,000 person threshold was reached, to ensure a sufficient population size to detect WNV at low incidence. Results were then downscaled back to the county level. Underlying spatial data from 2017 TIGER/Shapefiles from US Census bureau (US Census Bureau, 2021).

When risk was divided up into 5 equal intervals, 223 counties had the highest temperature-based risk for WNV, while 14 were in the lowest risk category (Fig. 3a). Most counties (84%, 365 of 433) were predicted to increase in temperature-based risk over the next decade (Fig. 3b).

**Fig. 3.**
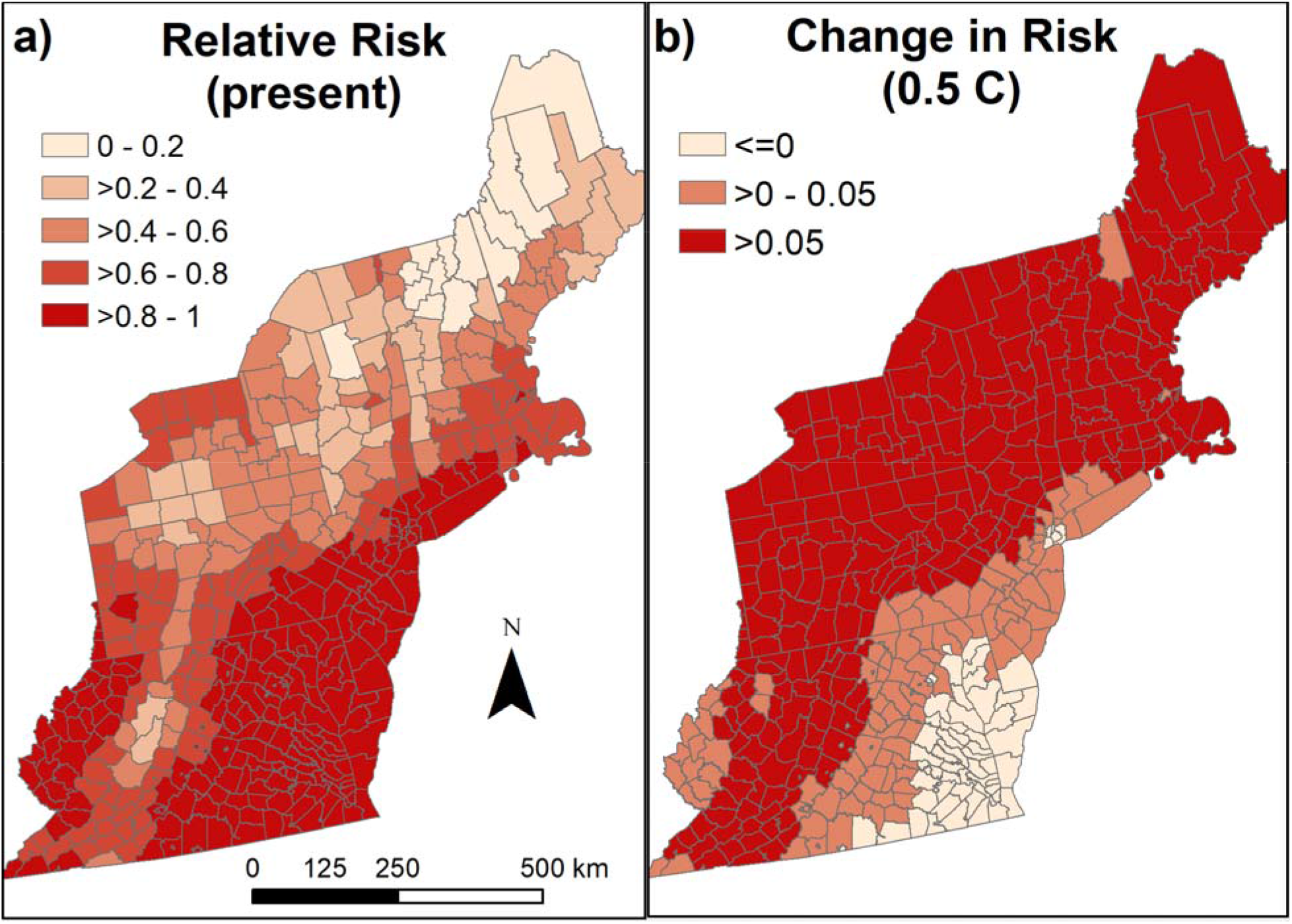
a) present-day temperature-trait-predicted relative risk for *Culex pipiens*, and b) the predicted change in temperature-based risk with 0.5 °C warming (on the high end for predicted for warming in the next decade). Note that this amount of warming falls within the range of present day temperature variation around the mean presented in (a). Map base layer from 2017 TIGER/Shapefiles (US Census Bureau, 2021).

The five most densely populated urban areas are expected to remain in a suitable temperature range for WNV under future warming (Table 1). Once 0.5 °C of warming has occurred, 4 of the 5 urban areas will decrease in risk with further warming. However, substantial reductions in risk for many major urban areas will not occur with less than 2.5 °C of warming.

## Discussion

A negative binomial distribution does a very good job of describing patterns of WNV in the northeast (Keyel et al., unpublished). The most important insight for public health is that a series of years with no or few WNV cases is possible even with a constant WNV risk. This means that reducing public health expenditures based on a few years with low WNV, on the assumption that it ‘has gone away’ is a poor strategy and will leave public health unprepared for the expected high years. WNV budgets should consider WNV risk over at least 5-year time horizons and have an emergency fund or the capacity to roll over funds from one year to the next, in order to address the expected high WNV years.

Further, areas with low rates of WNV may want to adopt a regional response approach that ensures counties have access to resources when cases occur. For most of the Northeast, a previous absence of a neuroinvasive WNV case over the past 20 years is not an indicator that the county will remain free of neuroinvasive WNV cases in the future, or even is lower risk than counties that have previously had cases. This suggests that these counties do not have some special protecting factors, but simply did not have cases due to low populations and random chance. The exceptions to this are in most of Maine (Fig. 2) and southwestern Virginia, where no cases have been reported. Both of these areas are predicted to have increased risk in the next decade due to climate change (Fig. 3). This is likely to be more relevant for Maine than Virginia, as current temperature-based risk is relatively low in Maine. In Virginia, temperature-based risk is already high, suggesting some other factor is responsible for the reduced number of cases. Therefore, southwestern Virginia may not see an increase in number of cases due to warming.

In the long-run, WNV risk is expected to increase across most of the Northeast, with the largest increases predicted in areas with relatively low present-day risk. Decreases in risk are predicted for the southern portion of the region. Locations where West Nile virus is relatively rare will need to be on the look-out for an increase in cases (Fig. 3b). These counties can expect to see substantial increases in temperature-based risk in the coming decades. Some of these regions should prepare to begin surveillance programs, doctors should familiarize themselves with WNV symptoms and lab work, and mosquito control operations should be prepared for expanded operations to reduce disease risk. That said, temperature-based risk is currently high in some localities that have low observed WNV risk, and therefore other factors may also be critical in determining how WNV risk may change into the future.

Locations with the most WNV cases in the present will have relatively little to do for long-term climate-change-related planning for West Nile virus. Existing mitigation measures should be as effective or more effective at controlling West Nile virus in the future, as conditions shift to be less suitable for mosquito-based transmission of West Nile virus. An important caveat is this research was purely from the standpoint of WNV. In locations where WNV is expected to decline, other vector-borne diseases, such as Zika virus may expand (Ryan et al., 2021). However, warming is not predicted to be sufficient to be suitable for dengue to become endemic in the Northeast under future climate change scenarios that extend out to 2080 (Messina et al., 2019).

The mismatch between probabilistic present-day risk compared to temperature-trait-based risk for the Northeast is interesting (compare Fig. 2b and Fig. 3a). Virginia and West Virginia are lower risk than expected based on the temperature models, while western and central Pennsylvania appears to be higher risk than predicted by the temperature models. Future work can explore whether landcover can explain these discrepancies, as prior research has suggested that urban areas are more favorable to WNV amplification (Bradley et al., 2008).

Another interesting future direction would be to compare probabilistic risk with present-day surveillance effort. Are some regions under-surveyed for vector-borne diseases? Could additional surveillance reduce human cases in these regions? Do some areas with high surveillance have fewer cases than predicted? Are they over-surveyed, or does the enhanced surveillance lead to fewer cases?

## Supporting information

STROBE Checklist

## Data Availability

The input data set is available from the United States Centers for Disease Control Division of Vector-Borne Diseases. Model outputs are available upon reasonable request to the authors.

## Acknowledgements

I thank A. Tyre for statistical assistance and O. Elison Timm for feedback on an earlier draft of the manuscript. I thank S. Mathis and the Centers for Disease Control for access to the CDC neuroinvasive data. This publication was supported by cooperative agreement 1U01CK000509-01, funded by the Centers for Disease Control and Prevention and by the National Institutes of Health grant R01AI168097. Its contents are solely the responsibility of the author and do not necessarily represent the official views of the Centers for Disease Control and Prevention or the Department of Health and Human Services.

---

**Figure S1.**
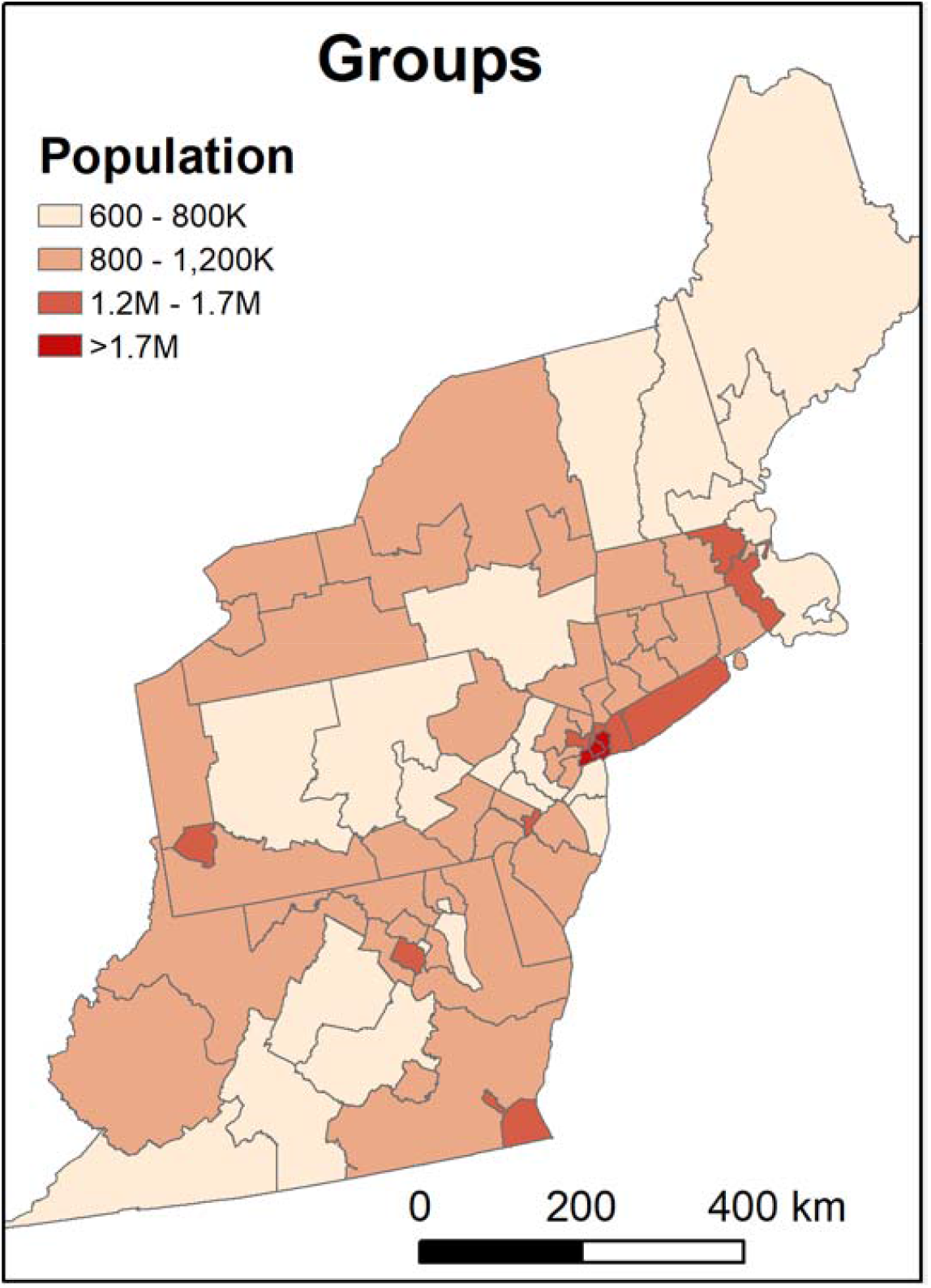
Map of population by groups used to fit the negative binomial model. Group assignment included a subjective element, and results for individual counties could vary depending on group assignment. Groups derived from merging 2017 TIGER/Shapefiles (US Census Bureau, 2021).

